# PROFILING THE IMMUNE RESPONSE TO THE DUFFY BINDING-LIKE 5 (DBL5) DOMAIN OF VAR2CSA IN PREGNANCY-ASSOCIATED MALARIA AMONG WOMEN IN NORTHERN GHANA

**DOI:** 10.1101/2025.09.29.25336887

**Authors:** Richmond Balinia Adda, Abdul-Rashid Mohammed, Jonah Kulariba, James Abugri

## Abstract

**Background:** Pregnancy-associated malaria (PAM), caused by *Plasmodium falciparum*, remains a major cause of maternal and neonatal mortality. The sequestration of infected erythrocytes in the placenta is mediated by the VAR2CSA protein. However, its high polymorphism complicates vaccine development. Targeting conserved sub-units like the Duffy Binding-Like 5 (DBL5) domain offers a potential alternative strategy. This study aimed to profile the immune response to the DBL5 domain and evaluate its cross-reactivity with VAR2CSA among pregnant women in Northern Ghana.

**Methods:** A subset of 156 plasma samples was selected from a cross-sectional study of pregnant women, categorized into *P. falciparum*-infected, hepatitis B-infected, and co-infected groups. The DBL5 domain was recombinantly expressed in *E. coli*, and its immunogenicity was assessed using indirect ELISA. Immunoinformatics tools were used to predict conserved B-cell and T-cell epitopes.

**Results:** ELISA results demonstrated significantly higher antibody reactivity to DBL5 in the *P. falciparum*-infected group (mean OD: 1.56 ± 0.12) compared to the hepatitis B-only (0.89 ± 0.15) and co-infection (1.22 ± 0.18) groups (p < 0.001). This indicates a specific and robust immune response to DBL5 in malaria-exposed women. Furthermore, plasma samples showed overlapping reactivity to both DBL5 and VAR2CSA, suggesting immunogenic cross-reactivity. Epitope mapping identified conserved DBL5 epitopes with broad global population coverage (>95%).

**Conclusion:** The DBL5 domain is highly immunogenic and demonstrates partial cross-reactivity with VAR2CSA. Our findings advocate for DBL5 as a promising, conserved candidate for inclusion in a multi-epitope vaccine against pregnancy-associated malaria.

## Background

Pregnancy-associated malaria (PAM) caused by *Plasmodium falciparum* is a severe public health challenge, particularly in sub-Saharan Africa, where malaria is endemic.^1^□Pregnant women, especially primigravidae, are at increased risk due to their lack of pre-existing immunity to the parasite.^2^□The sequestration of *Plasmodium falciparum*-infected erythrocytes in the placenta characterizes PAM, facilitated by the binding of the parasite’s Variant Surface Antigen 2-Chondroitin Sulphate A (VAR2CSA) to placental chondroitin sulphate A (CSA).^1^□,^22^ This interaction leads to inflammation, maternal anaemia, and poor pregnancy outcomes, including low birth weight and increased neonatal mortality.^2^,^1^□

Efforts to develop a vaccine against PAM have largely focused on VAR2CSA due to its critical role in mediating placental sequestration.□However, the antigenic diversity of VAR2CSA and the limited accessibility of conserved epitopes complicate vaccine design.^3^,□Alternative strategies involve targeting other domains within the *Plasmodium* genome that demonstrate immunogenic potential and cross-reactivity with VAR2CSA, thereby broadening the immune response against the parasite.□,□Among these alternative targets, the Duffy Binding-Like (DBL) proteins have garnered attention for their role in host-pathogen interactions.□DBL5, a subdomain of the VAR2CSA protein, has shown promise due to its ability to induce antibody responses in human plasma and its potential to elicit cross-reactive immunity.^13^,^2^□Studies indicate that recombinant DBL5 proteins can induce immunoglobulin G (IgG) antibodies capable of inhibiting CSA-binding, suggesting their suitability as a vaccine candidate.□,^1^□However, limited research has been conducted to assess its efficacy and cross-reactivity in diverse populations, particularly in malaria-endemic regions.^1^ This study seeks to profile the immune response to DBL5 in pregnant women in Northern Ghana, an area with a high prevalence of malaria during pregnancy.^1^□By examining the antigen-antibody interactions of DBL5 and its cross-reactivity with VAR2CSA, we aim to evaluate its potential as a conserved vaccine target.^1^□ Furthermore, we leverage immunoinformatics tools to identify conserved epitopes and assess population coverage to support the development of a vaccine with broad applicability.^1^□

## Materials and Methods

### Study Setting and Population

This study utilized a subset of samples and data from cross-sectional studies on hepatitis B and malaria conducted between October 2016 and February 2017 among pregnant women attending their first antenatal care (ANC) visit at selected health facilities in Northern Ghana. The Northern Region was selected due to its high ANC attendance rates. The specific study sites, chosen randomly from the districts, included Tamale West Hospital, Tamale Central Hospital, Bilpella Health Centre, Tamale Teaching Hospital, Sankpala Health Centre, and Kosawgu Health Centre (Figure 1).

**Figure 1:**
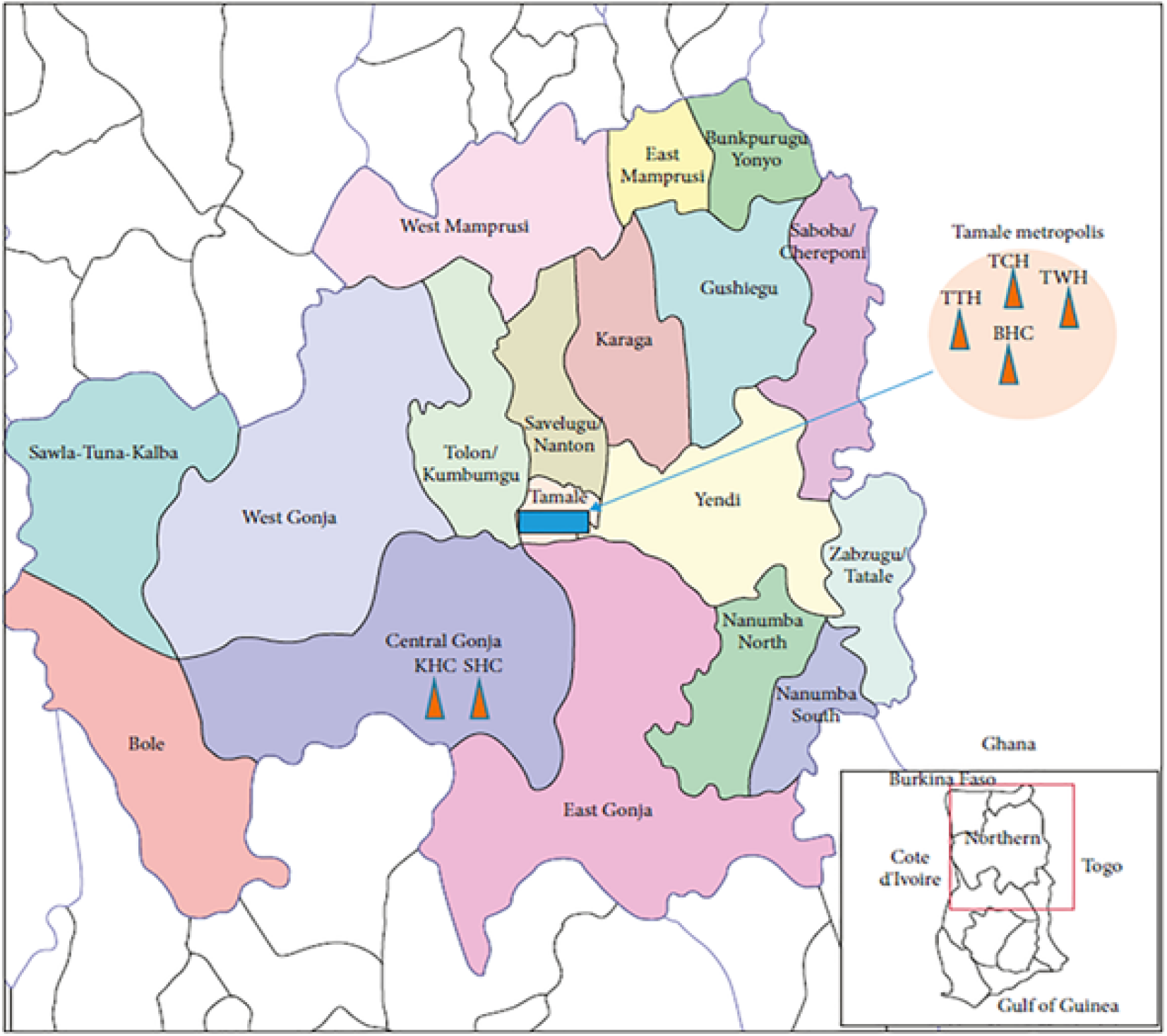
A map showing the locations of the research sites. TWH: Tamale West Hospital, TCH: Tamale Central Hospital, BHC: Bilpella Health Centre, TTH: Tamale Teaching Hospital, Sankpala Health Centre (SHC) and Kosawgu Health Centre (KHC)^1^

### Study Participants and Sample Selection

From the original cohort of 2,258 enrolled pregnant women, 187 participants were excluded due to incomplete clinical records or insufficient sample quality (e.g., hemolyzed plasma), resulting in 2,071 samples analyzed for infections and co-infections, as previously described (Anabire et al., 2019). For the present immunogenicity study, a final subset of 156 plasma samples was selected based on the following criteria:

1. Confirmed Plasmodium falciparum infection, determined by PCR or microscopy.
2. For the co-infection group, positivity for hepatitis B surface antigen (HBsAg).
3. Availability of complete obstetric and demographic records.

The participants were categorized into three groups: Malaria-only (n=60), Hepatitis B-only (n=60), and Co-infection (Malaria and Hepatitis B) (n=36). A flowchart detailing participant selection and group allocation is provided in Figure 2.

**Figure 2:**
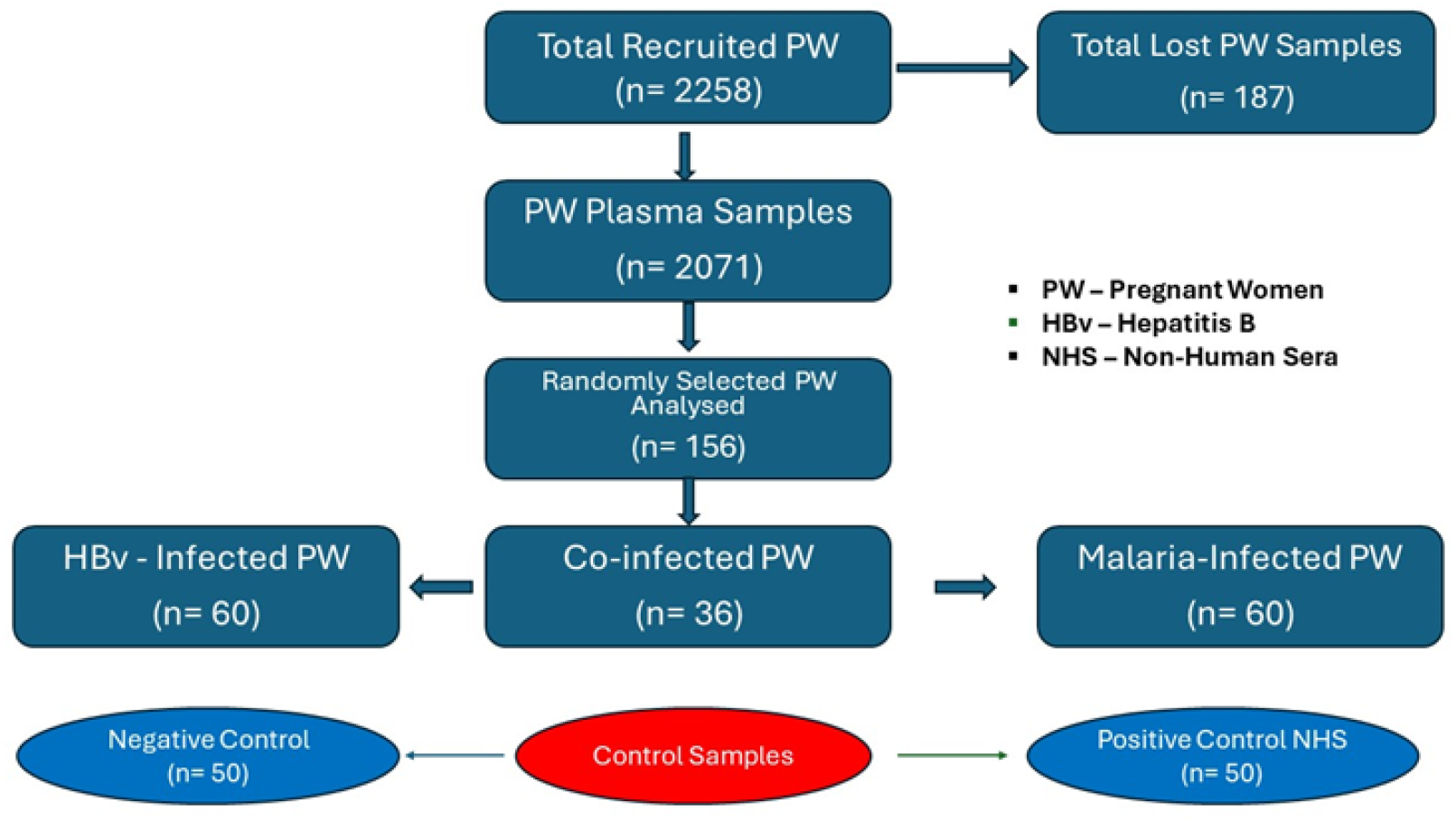
flowchart showing the clusters and groupings of study participants.

### Bioinformatic Analysis of DBL5

The protein sequence for the Duffy Binding-Like 5 (DBL5) domain (PlasmoDB ID: PF3D7_1335100) was obtained in FASTA format. Immunoinformatics analyses were performed to predict immunogenic epitopes. Linear B-cell epitopes were predicted using the BepiPred 2.0 algorithm within the Immune Epitope Database (IEDB), with a threshold set at 0.5. Cytotoxic T-cell (MHC-I) epitopes were predicted using the NetMHCpan 4.1 EL server. The population coverage of the predicted MHC-I epitopes was calculated using the IEDB population coverage calculation tool to assess potential global applicability.

### Recombinant DBL5 Protein Expression and Purification

The DBL5 gene was codon-optimized for expression in Escherichia coli and synthesized. The gene was cloned into a pET-based expression vector, incorporating an N-terminal hexahistidine (His-tag) tag. The recombinant plasmid was transformed into E. coli BL21(DE3) cells for protein expression.

Protein expression was induced in a 2 L culture with 1 mM isopropyl β-D-1-thiogalactopyranoside (IPTG) at 37°C for 4 hours. Bacterial cells were harvested by centrifugation, and the pellet was resuspended in lysis buffer (50 mM Tris-HCl, 300 mM NaCl, 10 mM imidazole, pH 8.0) supplemented with a protease inhibitor cocktail. The cells were lysed by sonication on ice, and the lysate was centrifuged at 9,000 × g for 45 minutes at 4°C to separate the soluble fraction.

The recombinant His-tagged DBL5 protein was purified from the soluble supernatant using immobilised metal affinity chromatography (IMAC) with Ni-NTA resin. The resin was washed with a buffer containing 50 mM imidazole to remove weakly bound contaminants, and the target protein was eluted with elution buffer containing 250 mM imidazole. The eluted protein was dialyzed against phosphate-buffered saline (PBS) to remove imidazole. Protein purity and molecular weight were confirmed by SDS-PAGE, and identity was verified by Western blotting using an anti-His tag antibody.

### Assessment of Antigen-Antibody Reactivity by Indirect ELISA

The immunoreactivity of plasma antibodies against the recombinant DBL5 protein was evaluated by indirect ELISA. High-binding 96-well plates (NUNC Maxisorp) were coated with 100 µl per well of purified DBL5 protein (2.0 µg/mL in PBS) and incubated overnight at 4°C. The plates were then blocked with 200 µl of blocking buffer (5% skim milk in PBS) for 2 hours at room temperature.

After blocking, plates were washed three times with PBS containing 0.05% Tween-20 (PBS-T). Subsequently, 100 µl of plasma samples, diluted 1:1000 in blocking buffer, were added to duplicate wells and incubated for 2 hours at room temperature. Following another wash step, 100 µl of horseradish peroxidase (HRP)-conjugated goat anti-human IgG secondary antibody, diluted 1:10,000, was added and incubated for 1 hour.

After a final wash, the reaction was developed by adding 100 µl of TMB Plus substrate and incubating in the dark for 20 minutes. The enzymatic reaction was stopped with 25 µl of 1 M H□SO□, and the optical density (OD) was measured at 450 nm using a microplate reader (Varioskan Lux). Each assay included positive control samples (from known malaria-infected individuals), negative controls (from healthy individuals), and blank wells (no plasma) for background subtraction.

### Statistical Analysis

Descriptive statistics were calculated for participant demographics and ELISA results. Standard Deviation (SD) and Standard Error of Mean (SEM) were used to determine the association between VAR2CSA-DBL5 and *Plasmodium falciparum*-infected placenta. Differences in optical density (OD) values across the three study groups (Malaria-only, Hepatitis B-only, and Co-infection) were assessed using a one-way Analysis of Variance (ANOVA). Following a significant ANOVA result, pairwise comparisons were conducted using Tukey’s post-hoc test. A p-value of <0.05 was considered statistically significant. Data analysis was performed using GraphPad Prism software (version 8.0).

### Ethical Considerations

The original studies and the use of stored samples for this analysis were approved by the Tamale Teaching Hospital Ethics Committee (Authorization Number: TTHERC/21/04/16/02). Written informed consent was obtained from all participants prior to their inclusion.

### Data Quality Assurance

All laboratory procedures, including protein purifications and ELISA, were performed in triplicate to ensure technical reproducibility. Negative and positive controls were included in every ELISA plate to validate assay performance and results.

## Results

### Participant Demographics

From the initial cohort of 2,071 pregnant women, a final subset of 156 participants was selected for this immunogenicity study. The demographic and obstetric profiles of the original cohort have been previously detailed (Anabire et al., 2019). The selected subset reflected a similar profile, with a mean age of 26.4 years (±4.5 SD). Over 60% of the participants were primigravidae, aligning with the study’s focus on populations at highest risk for pregnancy-associated malaria.

Expression and Purification of Recombinant DBL5 Protein The Duffy Binding-Like 5 (DBL5) domain was successfully expressed as a soluble, hexahistidine-tagged recombinant protein in *Escherichia coli* and purified using nickel-nitrilotriacetic acid (Ni-NTA) affinity chromatography. Analysis by SDS-PAGE revealed a predominant protein band at approximately 42 kDa (Figure 3A), which corresponds to the expected size of the DBL5 construct. The identity of the purified protein was confirmed by Western blot analysis using an anti-His tag antibody, which specifically detected the ∼42 kDa band (Figure 3B). The observed molecular weight is slightly higher than the theoretical weight of 38 kDa, a discrepancy potentially attributable to the physicochemical properties of the protein or minor post-translational modifications. The protein sequence, comprising 353 amino acids, was characterized bioinformatically, revealing an extinction coefficient of 77070 M□^1^ cm□^1^.

**Figure 3:**
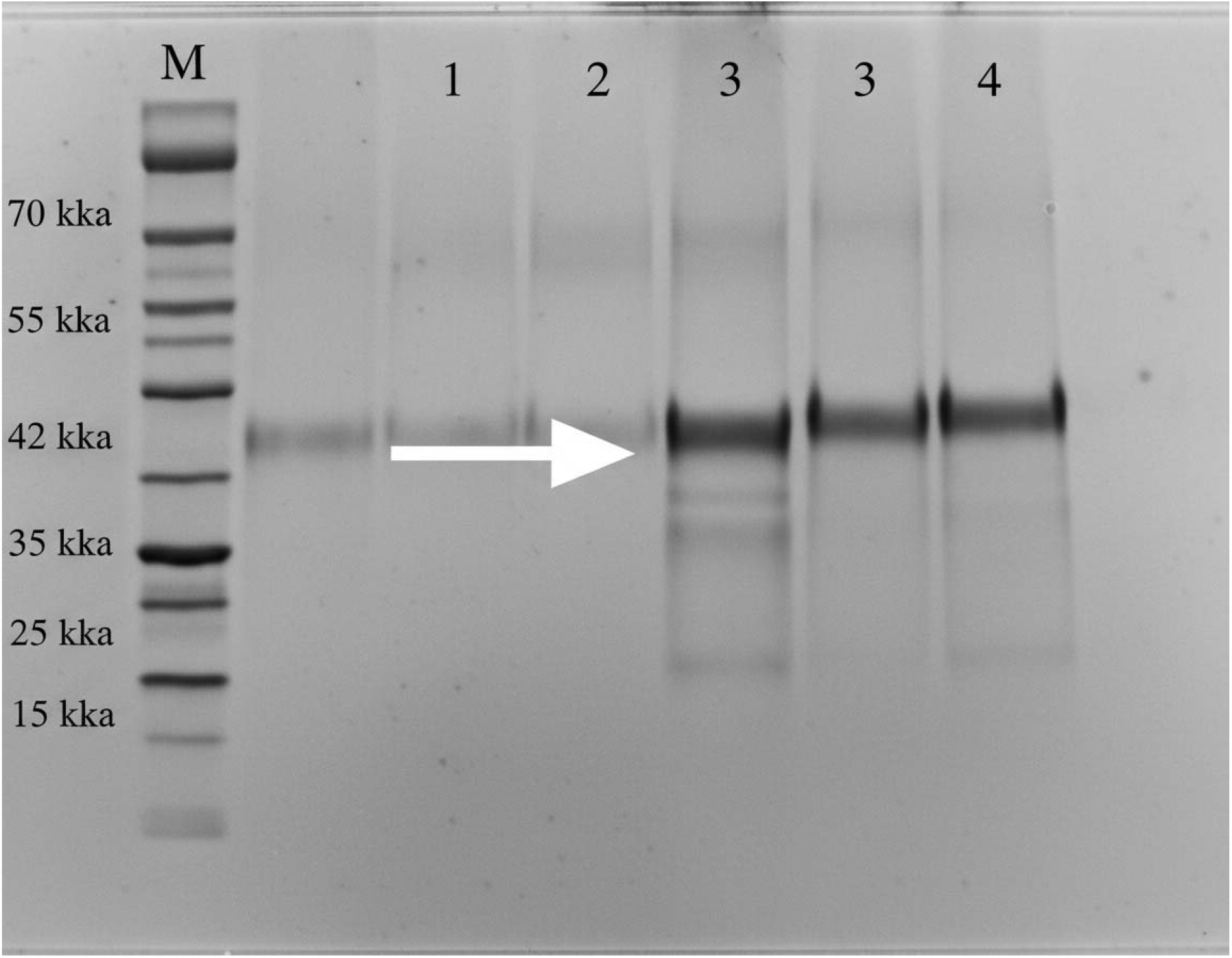
Characterization of recombinant DBL5 protein. (A) SDS-PAGE analysis of purified DBL5 showing a prominent band at ∼42 kDa. M: Protein molecular weight marker. (B) Western blot analysis of the same purification fraction, probed with an anti-His tag antibody, confirming the identity of the recombinant protein.

#### Segmented Sequence

1–60:

RCFDDQTKMK VCDLIGDAIG CKDKTKLDEL DEWNDMDLRD PYNKYKGVLI PPRRRQLCFS

61–120:

RIVRGPANLR NLNEFKEEIL KGAQSEGKFL GNYYNEDKDK EKKEDRKEKA LEAMKNSFYD

121–180:

YEYIIKGSDI LENIQFKDIK RKLDKLLTKE TNNNTKKAED WWETNKKSIW NAMLCGYKKS

181–240:

GNKIIDPSWC TIPTTEKTPQ FLRWIKEWGT NVCIQKEKYK EYVKSECSNV PNNNLGSQAS

241–300:

ESTKCTSEIR KYQEWSRKRS IQWEAISERY KKYKGMDEFK NVFNNANEPD ANEYLKEHCS

301–353:

KCPCGFNDME EITKYTNIGN EAFNTIIEKV KIPAELEDVI YRLKHHEYNS NDY

#### Plasma IgG Reactivity to DBL5 is Highest in Malaria-Infected Pregnant Women

The immunoreactivity of plasma IgG antibodies towards the recombinant DBL5 protein was quantitatively assessed by indirect ELISA. Optical density (OD) measurements at 450 nm revealed significant differences in antibody binding among the study groups (p < 0.001, one-way ANOVA).

As shown in Figures 4 and 5, plasma samples from the Malaria-only group exhibited the strongest reactivity, with a mean OD of 1.56 ± 0.12 (SEM). In contrast, samples from the Hepatitis B-only group showed significantly lower reactivity (mean OD: 0.89 ± 0.15), comparable to background levels. The Co-infection group (Malaria and Hepatitis B) demonstrated an intermediate level of antibody binding, with a mean OD of 1.22 ± 0.18. Post-hoc analysis confirmed that the OD values of the Malaria-only group were significantly higher than those of both the Hepatitis B-only and Co-infection groups.

**Figure 4:**
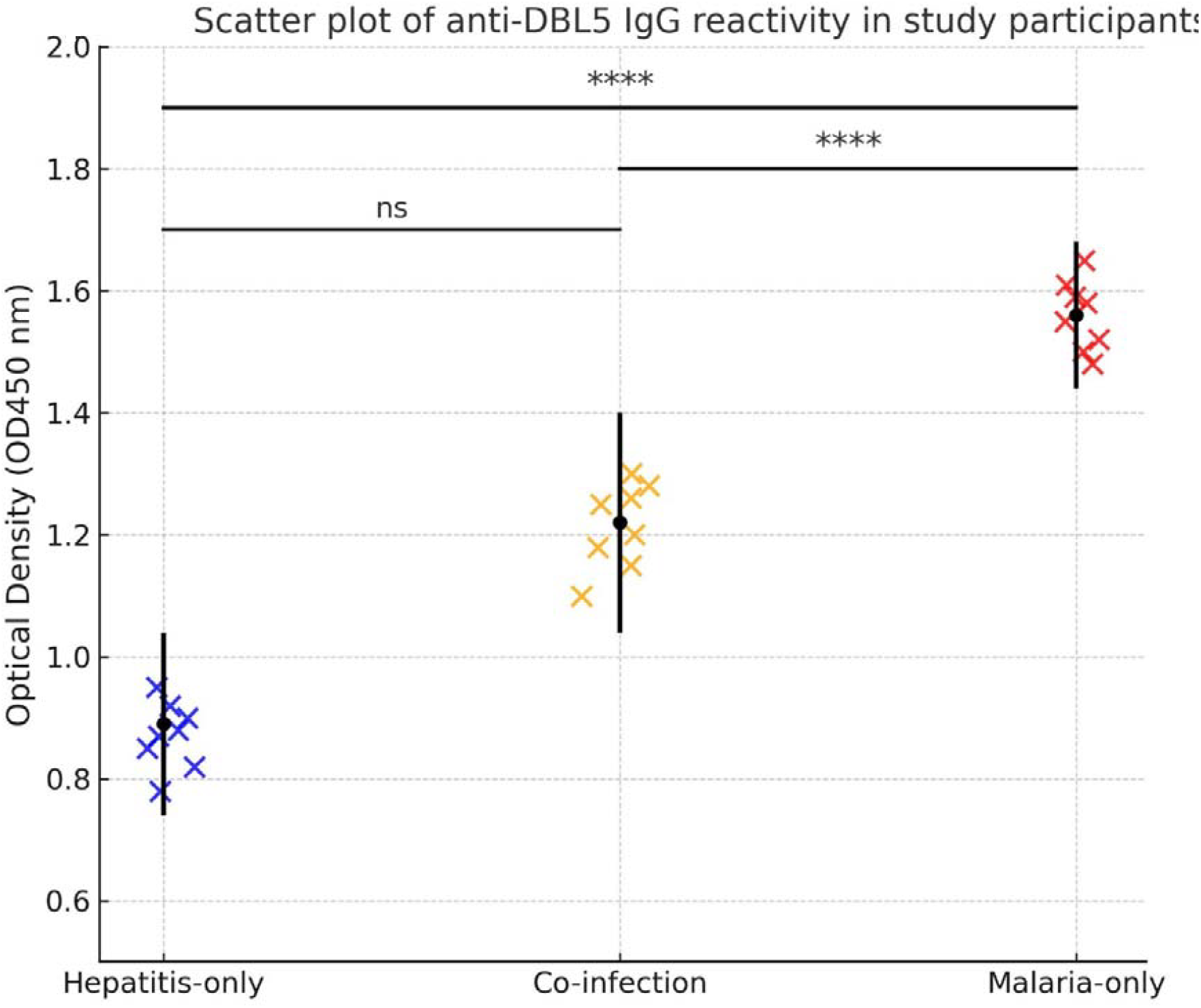
Scatter plot of anti-DBL5 IgG reactivity. Optical density (OD450) values for individual plasma samples from pregnant women with Malaria-only, Hepatitis B-only, or Co-infection. Horizontal bars represent the group mean. **** denotes p < 0.0001.

**Figure 5:**
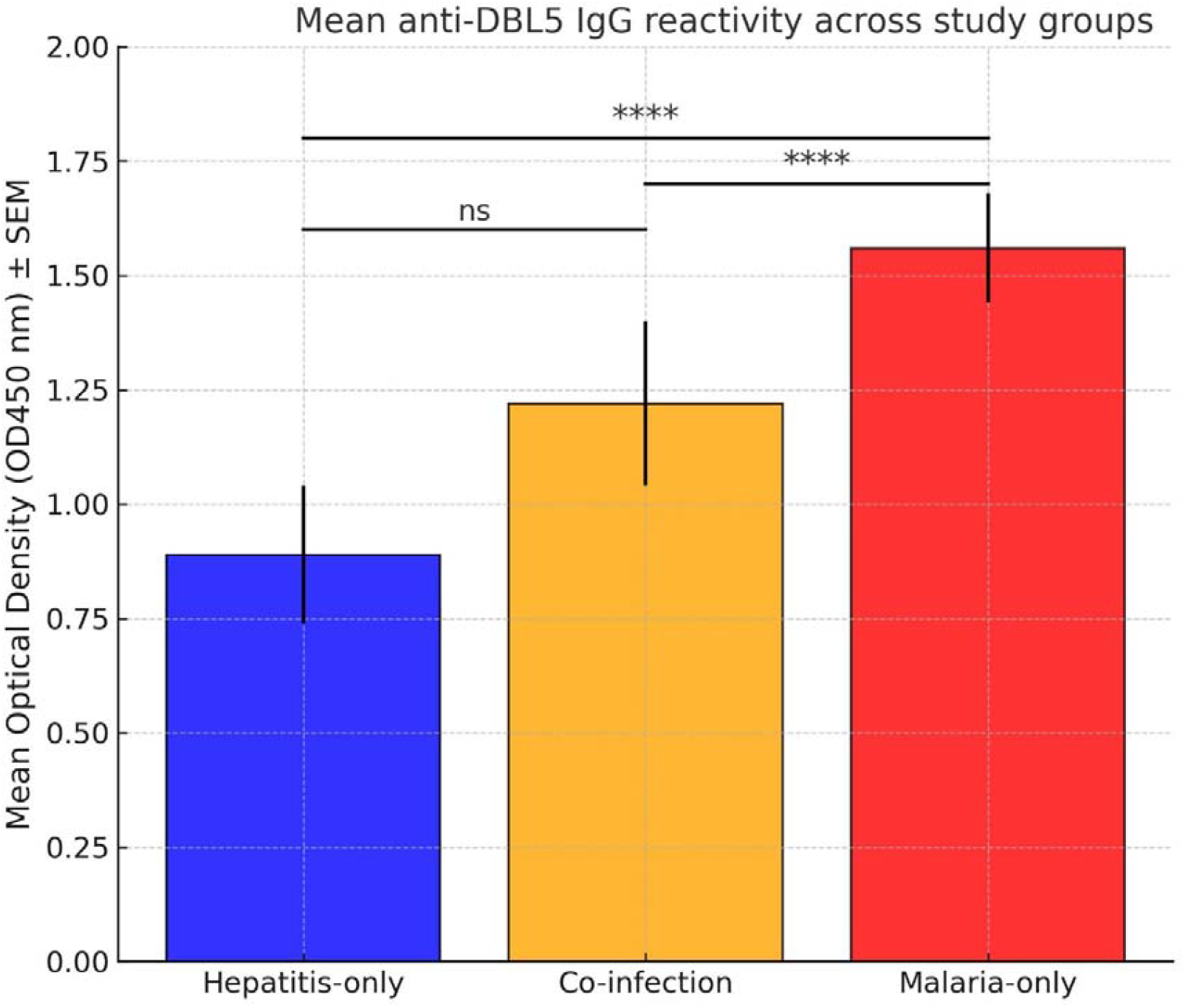
Bar graph of mean anti-DBL5 IgG reactivity. Data are presented as mean OD450 ± SEM for each study group. The Malaria-only group showed significantly higher antibody binding to DBL5 compared to the other groups.

#### Immunoinformatics Predicts Conserved B-cell and T-cell Epitopes in DBL5

In silico analysis was employed to predict potential immunogenic epitopes within the DBL5 sequence. B-cell epitope mapping using BepiPred 2.0 identified several linear epitopes distributed across the protein. Furthermore, analysis with NetMHCpan 4.1 EL predicted multiple high-affinity CD8+ T-cell (MHC-I) epitopes with strong binding affinities (IC50 < 200 nM) to common HLA alleles (Table 1). Notably, the global population coverage for the combined set of predicted T-cell epitopes was estimated to be greater than 95%, indicating their potential relevance across diverse genetic backgrounds, including those in malaria-endemic regions.

**Table 1:** Predicted high-affinity T-cell epitopes in the DBL5 protein.

| *Epitope Sequence* | *Start Position* | *End Position* | *Binding Affinity (IC50, nM)* | *MHC Allele* | *Global Population Coverage (%)* | *Comments* |
| --- | --- | --- | --- | --- | --- | --- |
| RCFDDQTKMK | 1 | 10 | 150 | HLA-A*02:01 | 96.5 | Conserved region; high affinity. |
| GNYYNEDKDK | 90 | 100 | 180 | HLA-B*07:02 | 95.8 | Overlaps with a predicted B-cell epitope. |
| TIPTTEKTPQ | 200 | 210 | 120 | HLA-A*11:01 | 98.0 | Strong T-cell epitope. |
| KYQEWSRKRS | 260 | 270 | 140 | HLA-A*24:02 | 97.2 | Cross-reactive potential. |
| KIPAELEDVI | 340 | 350 | 110 | HLA-B*44:02 | 99.1 | Highly conserved across strains. |

#### Evidence of Cross-Reactivity between DBL5 and VAR2CSA

To investigate the potential for shared immunity, we assessed the cross-reactivity of DBL5 with the full-length VAR2CSA protein. Comparative ELISA using a subset of plasma samples revealed overlapping binding patterns to both DBL5 and VAR2CSA. Sera with high reactivity to VAR2CSA also showed significant reactivity to DBL5, suggesting the presence of common epitopes recognized by naturally acquired antibodies. This indicates that DBL5 can elicit antibodies that cross-react with VAR2CSA, while also presenting unique immunogenic sites of its own.

## Discussion

This study’s findings provide vital insights into DBL5’s immunogenic potential as a target for pregnancy-associated malaria (PAM) vaccines, aligning with current efforts to improve maternal health outcomes in malaria-endemic regions.^12^ Our results highlight the relevance of targeting conserved epitopes within DBL5, as evidenced by its robust immune response and partial cross-reactivity with VAR2CSA.^13^

### Co-infection dynamics

The intermediate antigen-antibody reactivity observed in co-infected individuals (mean OD: 1.22 ± 0.18) suggests a complex interplay between malaria and hepatitis B immune responses. Hepatitis B virus (HBV)-induced immunosuppression or antigenic competition may modulate antibody production against DBL5, reducing cross-reactivity compared to malaria-only infections.^1^ Alternatively, HBV co-infection might amplify inflammatory pathways, indirectly enhancing anti-DBL5 responses. Further studies are needed to dissect these interactions and assess whether co-infections influence vaccine efficacy in endemic settings.

### Implications for vaccine design

While VAR2CSA remains the primary PAM vaccine candidate,^1^□its antigenic diversity poses a significant challenge.^3^,□Our study shows that DBL5’s distinct antigenic features— such as conserved epitopes with >95% global population coverage^1^□—complement VAR2CSA by broadening immune recognition. A multi-epitope vaccine combining DBL5 and VAR2CSA could target both conserved and variable regions, mitigating the impact of *Plasmodium falciparum*’s antigenic diversity.^1^□This strategy aligns with recent advances in monoclonal antibody therapies targeting conserved epitopes in VAR2CSA.^11^

### Comparative immunogenicity and epitope validation

Similar to our findings, subsequent investigations have highlighted VAR2CSA’s critical role in placental sequestration.^1^□Although vaccine candidates like PAMVAC and PRIMVAC have shown promising safety profiles, their efficacy across strains is limited due to antigenic variation.^12^,^1^□DBL5 provides a complementary advantage by retaining both unique and shared epitopes, potentially overcoming these restrictions. Its capacity to generate significant immune responses is consistent with observations that immunogens targeting conserved epitopes can boost cross-reactive immunity.^1^□Although immunoinformatics tools predicted high-affinity B-cell and T-cell epitopes in DBL5 (Table 1), experimental validation is crucial. Future work should employ peptide microarrays or ELISpot assays to confirm epitope immunogenicity and assess functional antibody responses (e.g., inhibition of CSA binding).^1^□

### Global population coverage and current trends

The substantial worldwide population coverage (>95%) projected for DBL5 epitopes^1^□supports its potential for broad-spectrum protection. This approach mirrors successful malaria vaccination efforts such as RTS,S/AS01 and R21/Matrix-M™, which target conserved regions of the circumsporozoite protein.^2^□Current PAM vaccination trials primarily focus on strain-specific protection, which may limit efficacy in diverse populations.^22^ Recent work on monoclonal antibodies targeting conserved VAR2CSA epitopes underscores the potential for broader protection through multi-antigen approaches.^11^

### Advances in immunoinformatics

The use of advanced immunoinformatics tools in this study follows current trends in epitope-based vaccine design.^21^ NetMHCpan and BepiPred, utilized here, have proven instrumental in recent vaccine trials, enabling precise predictions of T-cell and B-cell epitopes.^1^□Our findings indicate that DBL5, with its conserved and distinct antigenic features, could supplement VAR2CSA to enhance the breadth and durability of vaccine-induced immunity, offering a novel strategy to address PAM’s devastating impact on maternal and neonatal health.

### Limitations

While this study provides compelling evidence for DBL5 immunogenicity, more experimental confirmation of anticipated epitopes is required. Furthermore, large-scale trials with varied populations are necessary to validate the global applicability of DBL5-based therapies. Future studies should investigate multi-epitope vaccine formulations combining DBL5 and VAR2CSA to increase efficacy in preventing PAM.

## Conclusion

This study shows that the Duffy Binding-Like Protein 5 (DBL5) is a viable vaccination candidate for pregnancy-associated malaria (PAM). DBL5 generated robust immunological responses in plasma from malaria-infected pregnant women, demonstrating its immunogenic potential. Notably, the protein shows partial cross-reactivity with VAR2CSA, implying common epitopes while retaining distinct antigenic characteristics. This dual trait offers DBL5 as a complementary target to VAR2CSA, potentially increasing the breadth and efficacy of vaccine-induced immunity.

Furthermore, immunoinformatics investigations revealed conserved B-cell and T-cell epitopes within DBL5 with widespread global population coverage, notably in malaria-endemic areas. These findings emphasize DBL5’s ability to induce strain-transcending immunity, thereby resolving the issues faced by antigenic diversity in Plasmodium falciparum.

The findings of this study lay a solid platform for additional research into DBL5 in multi-epitope vaccine formulations, particularly for vulnerable populations like primigravid women. Future research should concentrate on experimental validation of anticipated epitopes, large-scale immunological studies, and the integration of DBL5 with other antigens to maximize vaccination efficacy. By pushing DBL5 as a novel target, our study adds to the global effort to address PAM’s devastating effects on maternal and neonatal health.

## Acknowledgements

We thank the staff of the West African Center for Biology of Infectious Pathogens (WACBIP) and the participating health facilities for their support. Special thanks to the study participants for their valuable contributions.

## Author Contributions

**Richmond Balinia Adda:** Conceptualisation, Methodology, Investigation, Formal Analysis, Data Curation, Writing – Original Draft Preparation, Visualisation.

**Abdul-Rashid Mohammed:** Methodology, Investigation, Writing – Review & Editing.

**Jonah Kulariba:** Methodology, Investigation, Writing – Review & Editing.

**James Abugri:** Conceptualisation, Writing – Review & Editing, Supervision, Project Administration.

## Data Availability

All data produced in the present study are available upon reasonable request to the authors

